# Impact of night shift work on vascular function in healthy adult workers

**DOI:** 10.64898/2025.12.04.25341671

**Authors:** Hiroyasu Murata, Ai Hirasawa, Ayumu Sakurai, Daichi Takano, Shotaro Saito, Marina Fukuie, Rina Suzuki, Masayoshi Horino, Shinya Matsushima, Shigeki Shibata

**Affiliations:** Department of Physical Therapy, Faculty of Health Science, Kyorin University, Mitaka, Tokyo, Japan; Department of Health and Welfare, Faculty of Health Sciences, Kyorin University, Mitaka, Tokyo, Japan; Department of Rehabilitation Science Specialization, Graduate School of Health Sciences, Kyorin University, Mitaka, Tokyo, Japan; Department of Biomedical Engineering, Toyo University, Kawagoe, Saitama, Japan; Department of General Medicine, Faculty of Medicine, Kyorin University, Mitaka City, Tokyo, Japan; Department of Trauma and Critical Care Medicine, Kyorin University, School of Medicine, Mitaka, Tokyo, Japan

**Keywords:** night shift work, vascular function, flow-mediated dilation, physical activity, sleep status

## Abstract

**Background:** Night shift work (NSW) is associated with increased cardiovascular risk, but its impact on vascular function and the role of lifestyle factors in healthy individuals remain unclear. We aimed to compare vascular function between NSW and day workers (DW) and to examine associations with objectively measured lifestyle behaviours.

**Methods:** In this cross-sectional study, 40 healthy adults (20 DW, 20 NSW) underwent assessments of flow-mediated dilation (FMD), brachial–ankle pulse wave velocity (baPWV), and ankle–brachial index (ABI). Physical activity and sleep status were objectively measured using triaxial accelerometry for 7 consecutive days. Group comparisons and correlation analyses were conducted to evaluate the effects of NSW and its association with lifestyle factors.

**Results:** FMD was significantly lower in the NSW group than in the DW group (4.0% vs. 7.0%, p < 0.01), while baPWV and ABI showed no between-group differences. In the overall cohort, FMD was positively correlated with time in bed, total sleep time, and sleep efficiency, and negatively correlated with nocturnal awakening. Subgroup analysis revealed that FMD was associated with sleep quality in DW and with physical activity patterns in NSW.

**Conclusions:** Chronic night shift work is associated with impaired endothelial function in healthy adults, potentially driven by disrupted sleep and reduced physical activity. Early assessment using FMD and targeted behavioral interventions may help preserve vascular health among shift workers.

**lay summary:** This study shows that healthy adults who regularly work night shifts have poorer blood vessel function than people who only work daytime hours, and that sleep and physical activity patterns may help explain this difference.

## INTRODUCTION

Night shift work (NSW) disrupts regular lifestyle patterns and has been associated with an increased risk of cardiovascular and metabolic diseases^1^. In particular, hypertension^2,3^, type 2 diabetes^4,5^, and obesity^6^—collectively referred to as lifestyle-related diseases—are closely linked to the progression of atherosclerosis and the onset of cardiovascular events^7,8^. Given the reported association between NSW and cardiovascular disease^1,9^, the early detection and prevention of vascular dysfunction in NSW workers is of growing importance.

Vascular function is commonly assessed using pulse wave velocity (PWV) and the ankle-brachial index (ABI) in clinical settings^10^. In addition, flow-mediated dilation (FMD), a non-invasive measure of endothelial function, has gained attention as an indicator of early atherosclerotic changes and a predictor of future cardiovascular events^11,12^. Although transient reductions in FMD immediately following NSW have been reported^13,14^, it remains unclear whether chronic NSW leads to sustained vascular impairment.

Lifestyle factors, such as physical activity and sleep status, are also essential for maintaining vascular health and may be adversely affected by NSW^15,16,17^. However, few studies have objectively assessed both vascular function and lifestyle behaviors concurrently among NSW workers.

We hypothesized that vascular function, as assessed by FMD, PWV, and ABI, is lower in NSW workers than in daytime workers and that this is associated with disruptions in lifestyle behaviors. Due to the limited existing evidence, this study was designed as an exploratory investigation. To test this hypothesis, we aimed to (1) compare vascular function indices between NSW and daytime workers, and (2) investigate the associations between vascular function and lifestyle factors such as physical activity and sleep status.

## MATERIALS AND METHODS

### Subjects

The participants were all volunteers. This study enrolled 40 healthy adult workers, comprising 20 individuals in the day work group (DW group) and 20 in the NSW group. The DW group included participants who had not engaged in NSW for at least the past year and were currently working only daytime shifts. The NSW group consisted of individuals who had worked night shifts at least once per week for at least the past year. Eligible participants were non-smokers, under 45 years of age, with a body mass index (BMI) below 30 kg/m^2^, and without a history of hypertension, dyslipidemia, or diabetes. Recruitment was conducted through public advertisements.

The study protocol was approved by the Ethics Committee of the Faculty of Health Sciences, Kyorin University (Approval No. 2022-61). All participants provided written informed consent after receiving a full explanation of the study procedures. The study was conducted in accordance with the Declaration of Helsinki.

### Study design

The study’s design was cross-sectional. All measurements were conducted in the morning to minimize circadian variation. Participants were instructed to avoid alcohol and caffeine, as well as vigorous physical activity, for 12 hours before testing, and to refrain from eating breakfast on the day of the assessment.

To control for the acute effects of NSW, data collection was avoided on the day following a night shift. It was scheduled on non-working days or on days before a scheduled night shift, ensuring that participants had adequate rest before testing.

### Sample size calculation

The required sample size was estimated assuming that a 3% difference in FMD would be clinically meaningful. This assumption was supported by two systematic reviews, which reported that each 1% decrease in brachial FMD is associated with a 12–13% increase in the risk of future cardiovascular events^18,19^. Therefore, a 3% difference in FMD corresponds to an approximate 36–39% increment in cardiovascular risk, which we considered clinically substantial. In addition, a previous study in a Japanese cohort reported a standard deviation of 2.9% for FMD^20^. Based on these values, a two-tailed independent samples t-test with α = 0.05 and power = 0.80 indicated that 15 participants per group were required. To account for variability and potential attrition, we planned to recruit at least 17 per group and ultimately enrolled 20 per group (total n = 40).

### Measurements

We assessed background information obtained from a self-administered questionnaire (including marital status, number of children, and menstrual cycle phase for female participants) and measured body composition and vascular function in a controlled laboratory setting using standardized procedures by trained personnel. In contrast, lifestyle factors, including physical activity and sleep status, were assessed over 7 days using validated self-monitoring tools in everyday conditions.

#### Body Composition

Body composition was assessed using a multi-frequency bioelectrical impedance analyzer (InBody® S10, InBody Japan Inc., Tokyo, Japan). Height and body weight were measured first, followed by the body composition assessment.

Participants removed their socks, and the electrode sites were cleaned with alcohol swabs to ensure good skin contact. Measurements were taken in a standing position with the arms abducted approximately 15° and the legs shoulder-width apart, ensuring that the arms and thighs did not touch the trunk or each other. Electrodes were attached to the thumbs and middle fingers of both hands, as well as to the ankles and heels of both feet.

#### Vascular Function

We evaluated vascular function using three indices: baPWV, ABI, and FMD. baPWV and ABI were measured using a vascular screening device (FORM-5, Fukuda Colin Co., Ltd., Tokyo, Japan) in a quiet, temperature-controlled room. Cuffs were placed on both upper arms and ankles, ECG electrodes were attached to the wrists, and a phonocardiograph microphone was placed on the sternum. Measurements were performed twice, and the average was used for analysis.

FMD was measured in the right brachial artery using an ultrasound diagnostic system (EPIQ 7, Philips, Eindhoven, The Netherlands) equipped with an 18 MHz linear-array probe, following the guidelines proposed by Thijssen et al^21^. After the arterial diameter was recorded for a 2-minute resting period, a forearm cuff (MDF® Bravata®, Germany) was inflated to 50 mmHg above the participant’s systolic blood pressure and maintained for 5 minutes. Upon cuff release, the arterial diameter was recorded for an additional 3 minutes.

Images were continuously captured at 30 Hz using a video capture card (Epiphan AV.io HD™, Canada) and stored on a hard drive. Arterial diameter was analysed using dedicated software (version 2.2.3, Takei Scientific Instruments Co., Japan), which automatically measured the distance between the anterior and posterior intima-media surfaces. Baseline diameter (D_base_) was defined as the average over the 2-minute resting period, and peak diameter (D_peak_) was automatically identified using a formerly reported algorithm^22^. FMD was calculated as the percent rise in peak from baseline diameter (D_base_) using the formula: FMD = [(D_peak_ − D_base_)/D_base_] × 100 (%). Measurements were performed twice with a 15-minute rest interval, and the average was used for analysis. Shear rate (SR) was calculated as 4 × blood flow velocity (cm/s) / vessel diameter (cm). The shear rate area under the curve (AUC) was assessed in two ways: (1) SR_peakAUC_, defined as the area from cuff release to the time of D_peak_, and (2) SR_30sAUC_, defined as the area from cuff release to 30 seconds thereafter. Both indices were calculated using the trapezoidal rule:Σ [½ (Xi+1 − Xi)(Yi+1 − Yi) + (Xi+1 − Xi)Yi], where X represents time and Y represents SR^23^. As FMD responses are influenced by shear stimulus, we calculated normalized FMD by dividing the FMD value by the corresponding SR_AUC_ value to account for differences in shear stimulus between individuals^24^.

#### Physical activity

Physical activity was measured using a triaxial accelerometer (wGT3X-BT, ActiGraph, Pensacola, FL, USA), a widely validated device in physical activity research^25,26^. Participants wore the device on their waists for 7 consecutive days during waking hours, excluding bathing or swimming. The device recorded raw acceleration data at a sampling rate of 30 Hz, with an epoch length set to 15 seconds^27^.

Data were analyzed using ActiLife software (ActiGraph, USA) to determine step counts, energy expenditure, and activity intensity levels. Based on the World Health Organization (WHO) guidelines, activity intensity was classified as sedentary (≤1.5 METs), light (1.6–2.9 METs), moderate (3.0–5.9 METs), or vigorous (≥6.0 METs)^28^.

The percentage of time spent in each intensity category was calculated, and daily averages from a seven-day monitoring period were used for subsequent analyses.

#### Sleep status

Sleep status was assessed using the same triaxial accelerometer (wGT3X-BT, ActiGraph, Pensacola, FL, USA), which was worn continuously at the waist for seven consecutive days, including during sleep. Sleep parameters were analyzed using ActiLife software (ActiGraph, USA), and sleep–wake periods were identified based on an algorithm proposed by Van Hees et al^29^.

The following sleep status variables were calculated: time in bed (TIB), total sleep time (TST), sleep efficiency, and wake after sleep onset (WASO). TIB was defined as the time from going to bed to getting out of bed, and TST was the total sleep duration scored during this period. Sleep efficiency was calculated as the percentage of TST relative to TIB. WASO was defined as the total duration of wakefulness after sleep onset. For all sleep variables, the average over the 7 days was used for analysis.

### Statistical analysis

Normality was assessed using the Shapiro–Wilk test. Variables with a normal distribution were analyzed using parametric tests, while those without a normal distribution were analyzed using non-parametric tests. Comparisons between the DW and NSW groups were conducted using unpaired t-tests or Mann–Whitney U tests for continuous variables, and chi-square tests for categorical variables.

Correlations between vascular function (FMD, baPWV, ABI) and lifestyle factors (physical activity, and sleep parameters) were assessed using Pearson’s or Spearman’s correlation coefficients, depending on data distribution. All results are presented as means ± standard deviations. A two-tailed p-value of <0.05 was considered statistically significant. All analyses were performed using EZR (Saitama Medical Center, Jichi Medical University, Saitama, Japan), a graphical user interface for R (The R Foundation for Statistical Computing, Vienna, Austria)^30^.

## RESULTS

### Participant characteristics and body composition

Participant characteristics are shown in Table 1. No significant differences were observed between the DW and NSW groups in age, sex, BMI, years of work experience, marital status, number of children, or menstrual cycle phase (all p > 0.05).

**Table 1.** Participant characteristics.

| Variable | DW group (n = 20) | NSW group (n = 20) | p-value |
| --- | --- | --- | --- |
| Age [years] | 30±5 | 31±4 | 0.25 |
| Sex [female/male] | 14 / 6 | 14 / 6 | 1.00 |
| BMI [kg/m<sup>2</sup>] | 20.3 (19.5–21.9) | 21.4 (20.2–23.8) | 0.13 |
| Work experience<br>[years] | 8 ± 3 | 10 ± 4 | 0.14 |
| Marital status<br>(married/unmarried) | 2 / 18 | 6 / 14 | 0.24 |
| Children (yes/no) | 1 / 19 | 3 / 17 | 0.61 |
| Menstrual cycle<br>(follicular/luteal) | 7 / 7 | 6 / 8 | 1.00 |
| Muscle mass [kg] | 37.0 (34.9–45.9) | 37.2 (34.7–41.0) | 0.99 |
| Body fat percentage<br>[%] | 21.0 ± 5.5 | 25.4 ± 6.3 | 0.02 |

Data are presented as mean ± standard deviation or median (interquartile range), as appropriate.
Abbreviations: BMI = body mass index; DW = day work; NSW = night shift work.

As shown in Table 1, the NSW group had a higher body fat percentage (21.0 ± 5.5% vs 25.4 ± 6.3%, p = 0.02) compared to the DW group. No differences were observed in muscle mass.

### Vascular function

Vascular function outcomes are shown in Table 2. No differences were observed in baPWV or ABI between the DW and NSW groups on either side (all p > 0.05). These results suggest that structural arterial stiffness and peripheral arterial status were comparable between the groups.

**Table 2.** Vascular function outcomes in DW and NSW groups.

| Variable | DW group (n = 20) | NSW group (n = 20) | p-value |
| --- | --- | --- | --- |
| Right baPWV [cm/s] | 1068 ± 109 | 1083 ± 132 | 0.70 |
| Left baPWV [cm/s] | 1073 ± 105 | 1083 ± 145 | 0.81 |
| Right ABI | 1.09 ± 0.06 | 1.10 ± 0.06 | 0.63 |
| Left ABI | 1.10 ± 0.08 | 1.09 ± 0.05 | 0.77 |

Data are presented as mean ± standard deviation.
Abbreviations: ABI = ankle–brachial index; baPWV = brachial–ankle pulse wave velocity; DW = day work; NSW = night shift work.

In contrast, FMD was lower in the NSW group compared to the DW group (7.0 ± 2.9% vs 4.0 ± 1.9%, p < 0.01), indicating reduced endothelial function in night shift workers. This difference remained after adjustment for shear rate. Specifically, FMD adjusted for SR_peakAUC_ was lower in the NSW group compared to the DW group (0.00038 ± 0.00015 vs 0.00022 ± 0.00011, p < 0.01), and similar results were obtained when adjusted for SR_30sAUC_ (0.00056 ± 0.00021 vs 0.00033 ± 0.00017, p < 0.01) (Figure 1).

**Figure 1.**
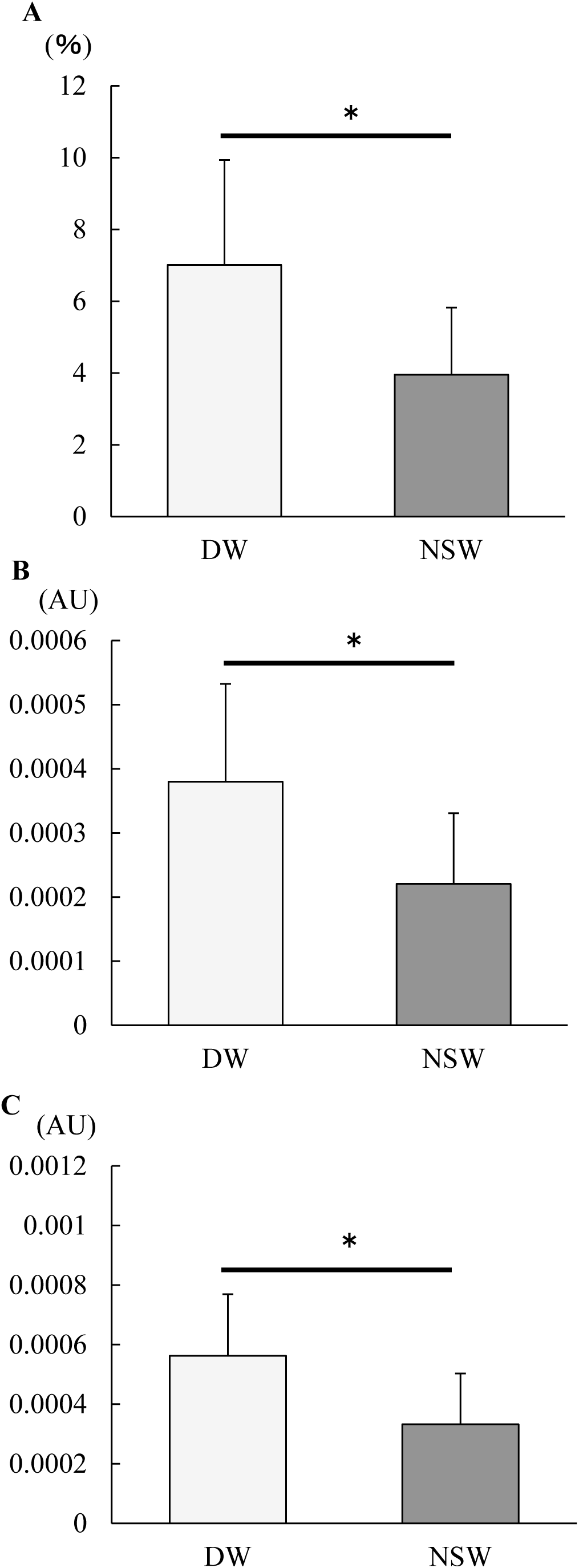
Endothelial function and related parameters between DW and NSW groups Values are shown as mean ± SD. *p < 0.01 A: FMD B: Normalized (SRpeakAUC) -FMD C: Normalized (SR30sAUC) -FMD Abbreviations: AU = arbitrary unit; DW = day work; FMD = flow-mediated dilation; NSW = night shift work; SRpeakAUC = shear rate peak area under the curve, SR30sAUC = shear rate 30 seconds area under the curve.

There were no differences between the two groups in any of the arterial variables measured during FMD, including baseline and peak diameter, baseline shear rate, SR_30sAUC_, SR_peakAUC_, and time to peak diameter (Table 3).

**Table 3.** Arterial variables during FMD measurements in DW and NSW groups.

| Variable | DW group (n = 20) | NSW group (n = 20) | p-value |
| --- | --- | --- | --- |
| D<sub>base</sub> [mm] | 3.51 ± 0.45 | 3.76 ± 0.62 | 0.15 |
| D<sub>peak</sub> [mm] | 3.75 ± 0.43 | 3.90 ± 0.59 | 0.34 |
| SR<sub>base</sub> [s<sup>-1</sup>] | 81.53 ± 27.77 | 81.27 ± 27.72 | 0.98 |
| SR<sub>peak</sub> [s<sup>-1</sup>] | 611.08 ± 152.25 | 608.65 ± 154.57 | 0.96 |
| SR<sub>30sAUC</sub> [AU] | 12592 ± 2875 | 12217 ± 2641 | 0.67 |
| SR<sub>peakAUC</sub> [AU] | 18832 ± 4106 | 18268 ± 2641 | 0.66 |
| Peak Time [sec] | 51.17 ± 11.05 | 51.13 ± 12.70 | 0.99 |

Data are presented as mean ± standard deviation.
Abbreviations: AU = arbitrary unit; D<sub>base</sub> = baseline diameter, D<sub>peak</sub> = peak diameter, DW = day work; FMD = flow-mediated dilation; NSW = night shift work; SR<sub>AUC</sub> = shear rate area under the curve, SR<sub>base</sub> = baseline shear rate, SR<sub>peakAUC</sub> = shear rate peak area under the curve, SR<sub>30sAUC</sub> = shear rate 30 seconds area under the curve, Peak Time = time to peak diameter from deflation.

### Lifestyle factors

Table 4 shows the physical activity results. The NSW group had a higher proportion of light-intensity activity than the DW group (13 ± 3% vs 16 ± 3%, p < 0.01). No differences were observed in energy expenditure, step counts, or other intensity levels.

**Table 4.** Physical activity levels in DW and NSW groups.

| Variable | DW (n = 20) | NSW (n = 20) | p-value |
| --- | --- | --- | --- |
| Energy expenditure [kcal] | 2,206 (1,495–3,098) | 2,284 (1,535–2,967) | 0.62 |
| Step counts [steps] | 70,465 (51,861–82,682) | 63,484 (55,221–79,766) | 0.94 |
| Sedentary [%] | 82 ± 4 | 80 ± 4 | 0.08 |
| Light [%] | 13 ± 3 | 16 ± 3 | <0.01 |
| Moderate [%] | 5 ± 2 | 4 ± 2 | 0.30 |
| Vigorous [%] | 0 ± 0 | 0 ± 0 | 0.18 |

Data are presented as mean ± standard deviation or median (interquartile range), as appropriate.
Abbreviations: DW = day work; NSW = night shift work.

Table 5 summarizes the sleep status. The NSW group showed shorter TIB (411 ± 37 vs 350 ± 45 min, p < 0.01) and TST (403 ± 38 vs 341 ± 46 min, p < 0.01) compared to the DW group. No differences were observed in either sleep efficiency or WASO.

**Table 5.** Sleep status in DW and NSW groups.

| Variable | DW group (n = 20) | NSW group (n = 20) | p-value |
| --- | --- | --- | --- |
| TIB [min] | 411 ± 37 | 350 ± 45 | < 0.01 |
| TST [min] | 403 ± 38 | 341 ± 46 | < 0.01 |
| Sleep efficiency [%] | 98 ± 1 | 97 ± 2 | 0.12 |
| WASO [min] | 7.81 ± 5.68 | 8.27 ± 3.99 | 0.77 |

Data are presented as mean ± standard deviation.
Abbreviations: DW = day work; NSW = night shift work; TIB = time in bed; TST = total sleep time; WASO = wake after sleep onset.

### Associations between FMD and lifestyle factors

No correlations were observed between FMD and physical activity variables in the overall cohort.

FMD showed associations with sleep-related parameters. FMD was positively correlated with TIB (r = 0.41, p < 0.01), TST (r = 0.44, p < 0.01), and sleep efficiency (r = 0.42, p < 0.01), and negatively correlated with WASO ( r = −0.34, p = 0.03), as shown in Figure 2.

**Figure 2.**
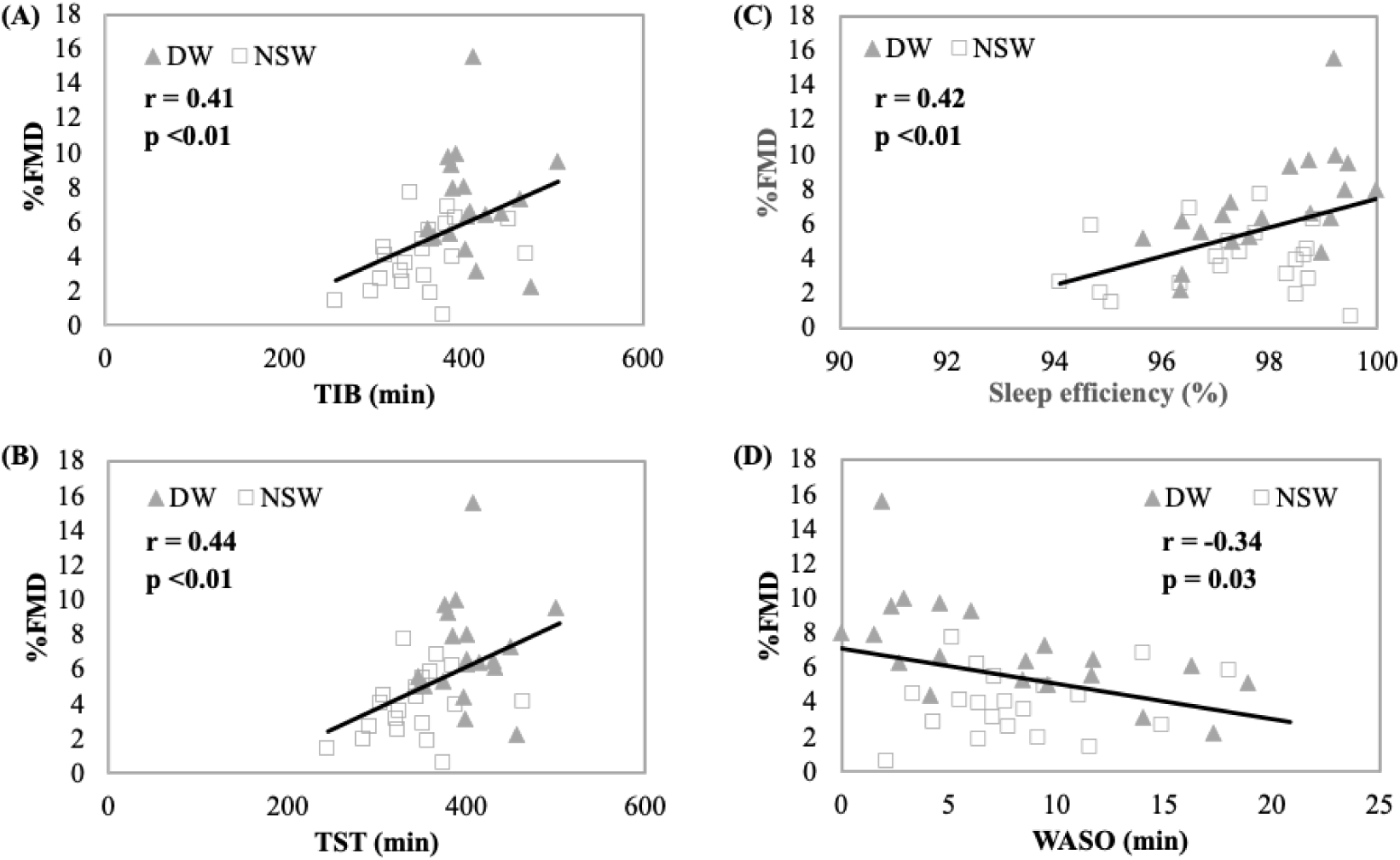
Relationships between %FMD and sleep parameters Scatter plots illustrate the relationships between %FMD and each sleep parameter: TIB, TST, sleep efficiency, and WASO. Grey triangles (▴) indicate participants in the DW group, and white squares (□) indicate those in the NSW group. The solid line represents the linear regression for all participants. A: Correlation between %FMD and TIB, B: Correlation between %FMD and sleep efficiency, C: Correlation between %FMD and TST, D: Correlation between %FMD and WASO. Abbreviations: DW = day work; FMD = flow-mediated dilation; NSW = night shift work; TIB = time in bed; TST = total sleep time; WASO = wake after sleep onset.

In subgroup analyses of the DW group, FMD was positively correlated with sleep efficiency (r = 0.62, p < 0.01) and negatively correlated with WASO (r = −0.64, p < 0.01), as shown in Figure 3. No correlations were observed between FMD and the physical activity parameters in the DW group.

**Figure 3.**
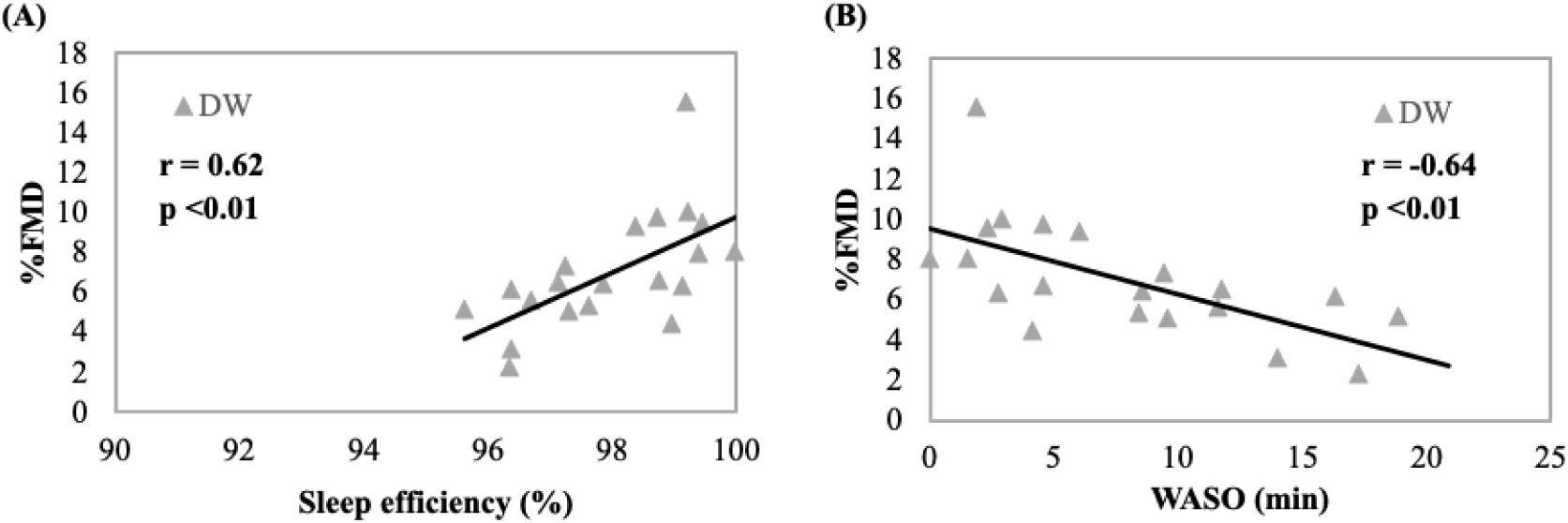
Relationships between %FMD and physical activity parameters in DW group Scatter plots illustrate the relationships between %FMD and each sleep parameter: sleep efficiency and WASO. A: Correlation between %FMD and sleep efficiency, B: Correlation between %FMD and WASO. Abbreviations: DW = day work; FMD = flow-mediated dilation; WASO = wake after sleep onset.

In the NSW group, FMD was negatively correlated with sedentary time (r = −0.47, p = 0.04) and positively correlated with time spent in light-intensity physical activity (r = 0.52, p = 0.02), as shown in Figure 4. No correlations were observed between FMD and sleep parameters in the NSW group.

**Figure 4.**
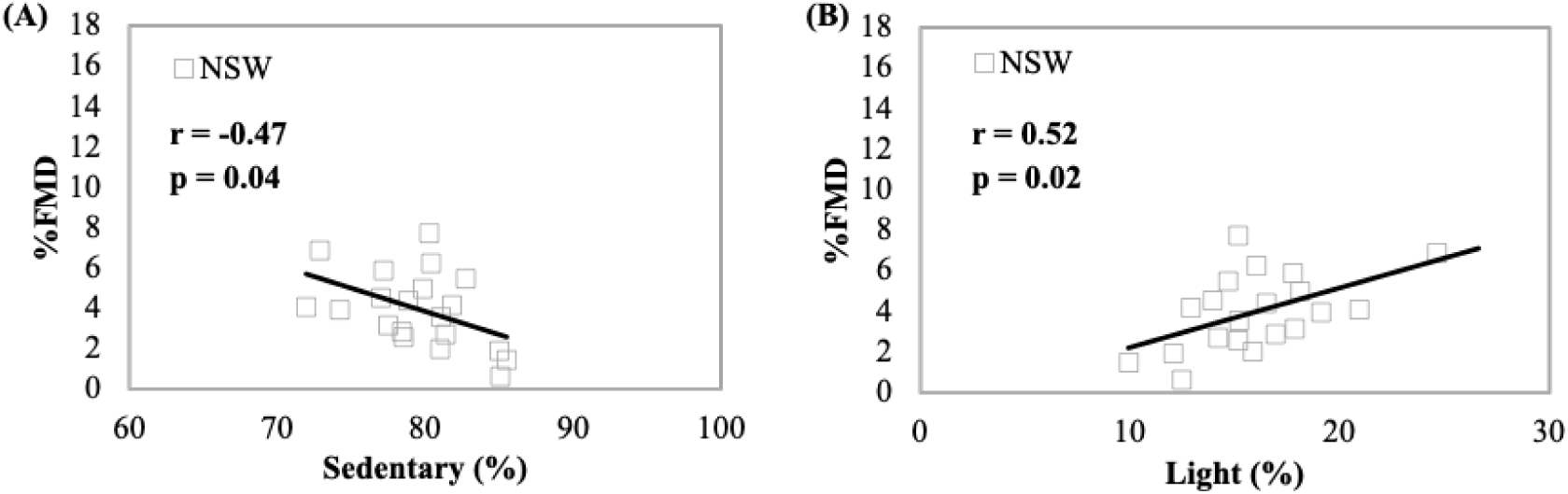
Relationships between %FMD and sleep parameters in NSW group Scatter plots illustrate the relationships between %FMD and each physical activity parameters: sedentary and light. A: Correlation between %FMD and rate of sedentary, B: Correlation between %FMD and rate of light intensity. Abbreviations: FMD = flow-mediated dilation; NSW = night shift work.

## DISCUSSION

### Summary of main findings

This cross-sectional study investigated the impact of night shift work on vascular function in healthy adult workers. The NSW group demonstrated lower FMD than the DW group, suggesting impaired endothelial function. However, there were no differences in baPWV or ABI between the groups. Regarding lifestyle factors, FMD was positively associated with sleep indicators (TIB, TST, and sleep efficiency) and negatively associated with WASO in the overall sample. Subsequent stratified analyses further revealed correlations between FMD and sleep efficiency, as well as between FMD and WASO, in the DW group. In contrast, the FMD was associated with light physical activity and sedentary time in the NSW group. These findings suggest that night shift work may impair endothelial function, and that this impairment is associated with lifestyle behaviors such as physical activity and sleep.

### Two-group comparison of vascular indices

Although FMD was lower in the NSW group, no differences in baPWV or ABI were observed between the groups. This finding indicates that while endothelial dysfunction may already be present, more advanced structural changes in the vasculature or indicators of peripheral artery disease may not yet have developed. FMD has been demonstrated to reflect nitric oxide (NO)-dependent vasodilation and is thus regarded as a sensitive marker of early-stage atherosclerosis^31^. In contrast, baPWV quantifies arterial stiffness, while ABI detects peripheral arterial stenosis or occlusion, both of which reflect more advanced structural vascular abnormalities. Therefore, the lack of alterations in baPWV or ABI, the diminished FMD, indicates that our metrics effectively discerned early-stage vascular dysfunction. Given the relatively young age of the cohort (approximately 30 years) and the exclusion of individuals with significant risk factors, such as obesity, hypertension, or diabetes, the absence of substantial changes in baPWV or ABI is consistent with expected findings. In contrast, FMD may be a more sensitive indicator for detecting subtle vascular impairment in younger NSW adults.

### Potential mechanisms linking NSW and FMD

Multiple factors have been implicated in the reduction of FMD, including aging, smoking, chronic inflammation, and sleep disturbances^13,14,32–36^. In the present study, in addition to the reduction in FMD among NSW participants, these individuals exhibited shorter TIB and TST and longer WASO. These findings suggest that insufficient and fragmented sleep may contribute to endothelial dysfunction. This finding aligns with the conclusions of previous studies, which have indicated that sleep deprivation impairs endothelium-dependent vasodilation^37,38^. Furthermore, the disruption of circadian rhythms induced by rotating shift schedules may also be a critical factor. Recent studies have demonstrated a correlation between circadian misalignment and immune cell dysfunction, as well as impaired inflammatory responses involved in endothelial regulation^39^. In addition, melatonin has been shown to modulate circadian rhythms and is closely linked to sleep^40^. Melatonin has also been reported to possess antioxidant effects^41^, and studies indicate it improves vascular endothelial function in patients with heart failure with reduced ejection fraction^42^. This finding may indicate a possible decrease in melatonin production in the NSW group. However, the present study did not investigate factors such as hormones, including melatonin, low-grade inflammation, or oxidative stress. This aspect warrants further investigation in subsequent research endeavors.

### The role of lifestyle factors in DW vs. NSW

In analyses stratified by group, the associations between FMD and sleep indicators were particularly pronounced in the DW group. In the overall sample, FMD demonstrated a moderate positive association with TIB, TST, and sleep efficiency, and a negative association with WASO. These findings suggest that both sufficient and consolidated sleep contribute to maintaining endothelial health. The present findings are consistent with the conclusions of previous studies, which reported that poor sleep quality impairs vascular reactivity^37,38^. Among subjects in the DW group, FMD showed a correlation with sleep efficiency and WASO. Conversely, among the NSW participants, several activity-related variables correlated with FMD, including sedentary time (negative correlation) and light physical activity (positive correlation). The present findings are consistent with earlier research, which demonstrated that prolonged sedentary behavior and insufficient physical activity are associated with lower FMD^43–45^. While previous studies have primarily focused on older adults or patients with chronic diseases, the present study demonstrates similar associations in young NSW workers, underscoring the importance of encouraging physical activity to preserve endothelial function in this population.

### Limitations and perspectives

The present study is not without its limitations. Firstly, due to its cross-sectional design, causal relationships between night shift work and vascular function cannot be firmly established. Future longitudinal studies are warranted to investigate temporal changes in vascular function in relation to shift frequency and cumulative shift work exposure. Secondly, given that the majority of participants in the NSW group were engaged in rotating shift schedules, it was challenging to discern the specific effects of night work per se from those of frequent schedule alternation. Furthermore, discrepancies in occupation or job responsibilities may have influenced both lifestyle behaviors and physiological outcomes. Thirdly, the present study was exploratory in nature, and the analyses were limited to descriptive and univariable approaches without multivariable adjustment. Consequently, residual confounding may persist, necessitating a circumspect interpretation of the observed associations.

Future research should include more detailed documentation of work patterns, as well as multidimensional assessments incorporating hormonal profiles and circadian rhythm biomarkers. Furthermore, interventional studies evaluating the effects of modifying physical activity or sleep behaviors on endothelial function may facilitate the development of effective prevention strategies for shift workers.

## CONCLUSION

In this study, we examined endothelial function in healthy young adult workers who worked either the night or the day shift. Our findings demonstrated that night shift workers exhibited significantly lower FMD values compared to day workers, suggesting impaired endothelial function. Conversely, baPWV and ABI showed no substantial differences between groups, suggesting that the observed endothelial dysfunction may represent an early stage of vascular impairment, preceding the onset of structural changes. Correlation analyses further revealed that, among night shift workers, FMD was associated with physical activity parameters. Conversely, among day workers, a stronger correlation with sleep quality was observed, suggesting that the factors influencing vascular health may vary by work schedule.

These findings underscore the importance of early assessment of endothelial function in night shift workers and support the need for preventive interventions targeting modifiable lifestyle factors.

## Data Availability

The datasets generated and analyzed during the current study are not publicly available due to ethical and privacy restrictions but are available from the corresponding author on reasonable request.

## Acknowledgements

We would like to express our sincere gratitude to all the participants in this study for their time, cooperation, and commitment. Without their invaluable contribution, this research would not have been possible.

## References

1. Vyas MV, Garg AX, Iansavichus AV, Costella J, Donner A, Laugsand LE, Janszky I, Mrkobrada M, Parraga G, Hackam DG. Shift work and vascular events: systematic review and meta-analysis. BMJ . 2012;345:e4800.

2. Oishi M, Suwazono Y, Sakata K, Okubo Y, Harada H, Kobayashi E, Uetani M, Nogawa K. A longitudinal study on the relationship between shift work and the progression of hypertension in male Japanese workers. Journal of hypertension. 2005;23(12):2173–2178.

3. Morikawa Y, Nakagawa H, Miura K, Ishizaki M, Tabata M, Nishijo M, Higashiguchi K, Yoshita K, Sagara T, Kido T, Naruse Y, Nogawa K. Relationship between shift work and onset of hypertension in a cohort of manual workers. Scandinavian journal of work, environment & health. 1999;25(2):100–104.

4. Pan A, Schernhammer ES, Sun Q, Hu FB. Rotating night shift work and risk of type 2 diabetes: two prospective cohort studies in women. PLoS medicine. 2011;8(12):e1001141.

5. Eriksson A-K, van den Donk M, Hilding A, Östenson C-G. Work stress, sense of coherence, and risk of type 2 diabetes in a prospective study of middle-aged Swedish men and women. Diabetes care. 2013;36(9):2683–2689.

6. Khosravipour M, Khanlari P, Khazaie S, Khosravipour H, Khazaie H. A systematic review and meta-analysis of the association between shift work and metabolic syndrome: The roles of sleep, gender, and type of shift work. Sleep medicine reviews. 2021;57(101427):101427.

7. van Trier TJ, Mohammadnia N, Snaterse M, Peters RJG, Jørstad HT, Bax WA. Lifestyle management to prevent atherosclerotic cardiovascular disease: evidence and challenges. Netherlands heart journal: monthly journal of the Netherlands Society of Cardiology and the Netherlands Heart Foundation. 2022;30(1):3–14.

8. Doughty KN, Del Pilar NX, Audette A, Katz DL. Lifestyle medicine and the management of cardiovascular disease. Current cardiology reports. 2017;19(11):116.

9. Kecklund G, Axelsson J. Health consequences of shift work and insufficient sleep. BMJ (Clinical research ed.). 2016;355:i5210.

10. Green DJ, Hopman MTE, Padilla J, Laughlin MH, Thijssen DHJ. Vascular adaptation to exercise in humans: Role of hemodynamic stimuli. Physiological reviews. 2017;97(2):495–528.

11. Perticone F, Ceravolo R, Pujia A, Ventura G, Iacopino S, Scozzafava A, Ferraro A, Chello M, Mastroroberto P, Verdecchia P, Schillaci G. Prognostic significance of endothelial dysfunction in hypertensive patients. Circulation. 2001;104(2):191–196.

12. Schächinger V, Britten MB, Zeiher AM. Prognostic impact of coronary vasodilator dysfunction on adverse long-term outcome of coronary heart disease. Circulation. 2000;101(16):1899–1906.

13. Tarzia P, Milo M, Di Franco A, Di Monaco A, Cosenza A, Laurito M, Lanza GA, Crea F. Effect of shift work on endothelial function in young cardiology trainees. European journal of preventive cardiology. 2012;19(5):908–913.

14. Zheng H, Patel M, Hryniewicz K, Katz S. Association of extended work shifts, vascular function, and inflammatory markers in internal medicine residents: a randomized crossover trial. JAMA. 2006;296(9):1049–1050.

15. Bouillon-Minois J-B, Thivel D, Croizier C, Ajebo É, Cambier S, Boudet G, Adeyemi OJ, Ugbolue UC, Bagheri R, Vallet GT, Schmidt J, Trousselard M, Dutheil F. The Negative Impact of Night Shifts on Diet in Emergency Healthcare Workers. Nutrients. 2022;14(4).

16. Hulsegge G, Proper KI, Loef B, Paagman H, Anema JR, van Mechelen W. The mediating role of lifestyle in the relationship between shift work, obesity and diabetes. International archives of occupational and environmental health. 2021;94(6):1287–1295.

17. Kim S-Y, Lee MY, Kim SI, Lim W-J. The mediating effects of working hours, sleep duration, and depressive mood on the association between shift work and the risk of suicidal ideation in Korean workers. Sleep medicine. 2022;93:49–55.

18. Matsuzawa Y, Kwon T-G, Lennon RJ, Lerman LO, Lerman A. Prognostic value of flow-mediated vasodilation in brachial artery and fingertip artery for cardiovascular events: A systematic review and meta-analysis. Journal of the American Heart Association. 2015;4(11).

19. Inaba Y, Chen JA, Bergmann SR. Prediction of future cardiovascular outcomes by flow-mediated vasodilatation of brachial artery: a meta-analysis. The international journal of cardiovascular imaging. 2010;26(6):631–640.

20. Shimada K, Fukuda S, Maeda K, Kawasaki T, Kono Y, Jissho S, Taguchi H, Yoshiyama M, Yoshikawa J. Aromatherapy alleviates endothelial dysfunction of medical staff after night-shift work: preliminary observations. Hypertension research: official journal of the Japanese Society of Hypertension. 2011;34(2):264–267.

21. Thijssen DHJ, Black MA, Pyke KE, Padilla J, Atkinson G, Harris RA, Parker B, Widlansky ME, Tschakovsky ME, Green DJ. Assessment of flow-mediated dilation in humans: a methodological and physiological guideline. American journal of physiology. Heart and circulatory physiology. 2011;300(1):H2–12.

22. Black MA, Cable NT, Thijssen DHJ, Green DJ. Importance of measuring the time course of flow-mediated dilatation in humans. Hypertension. 2008;51(2):203–210.

23. Kamoda T, Sakamoto R, Katayose M, Yamamoto S, Neki T, Sato K, Iwamoto E. Skipping breakfast does not accelerate the hyperglycemia-induced endothelial dysfunction but reduces blood flow of the brachial artery in young men. European journal of applied physiology. 2024;124(1):295–308.

24. Cowley AW Jr, Liard JF, Guyton AC. Role of the baroreceptor reflex in daily control of arterial blood pressure and other variables in dogs. Circulation research. 1973;32(5):564–576.

25. Vetrovsky T, Siranec M, Frybova T, Gant I, Svobodova I, Linhart A, Parenica J, Miklikova M, Sujakova L, Pospisil D, Pelouch R, Odrazkova D, Parizek P, Precek J, Hutyra M, et al. Lifestyle walking intervention for patients with heart failure with reduced ejection fraction: The WATCHFUL trial. Circulation. 2024;149(3):177–188.

26. Huang W-C, Chang S-H, Hsueh M-C, Liao Y. Relationship of sleep regularity with device-based sedentary behavior time and physical activity time in working adults. Sleep health. 2023;9(1):86–92.

27. Luo X, Herold F, Ludyga S, Gerber M, Kamijo K, Pontifex MB, Hillman CH, Alderman BL, Müller NG, Kramer AF, Ishihara T, Song W, Zou L. Association of physical activity and fitness with executive function among preschoolers. International journal of clinical and health psychology: IJCHP. 2023;23(4):100400.

28. Health Organization W. WHO guidelines on physical activity and sedentary behaviour. 2020.

29. van Hees VT, Sabia S, Anderson KN, Denton SJ, Oliver J, Catt M, Abell JG, Kivimäki M, Trenell MI, Singh-Manoux A. A novel, open access method to assess sleep duration using a wrist-worn accelerometer. PloS one. 2015;10(11):e0142533.

30. Kanda Y. Investigation of the freely available easy-to-use software “EZR” for medical statistics. Bone marrow transplantation. 2013;48(3):452–458.

31. Green DJ, Jones H, Thijssen D, Cable NT, Atkinson G. Flow-mediated dilation and cardiovascular event prediction: does nitric oxide matter?: Does nitric oxide matter? Hypertension. 2011;57(3):363–369.

32. Amir O, Alroy S, Schliamser JE, Asmir I, Shiran A, Flugelman MY, Halon DA, Lewis BS. Brachial artery endothelial function in residents and fellows working night shifts. The American journal of cardiology. 2004;93(7):947–949.

33. Wehrens SMT, Hampton SM, Skene DJ. Heart rate variability and endothelial function after sleep deprivation and recovery sleep among male shift and non-shift workers. Scandinavian journal of work, environment & health. 2012;38(2):171–181.

34. Seals DR, Jablonski KL, Donato AJ. Aging and vascular endothelial function in humans. Clinical science (London, England: 1979). 2011;120(9):357–375.

35. Jia X, Zhang P, Meng L, Tang W, Peng F. The association between smoking exposure and endothelial function evaluated using flow-mediated dilation values: a meta-analysis. BMC cardiovascular disorders. 2024;24(1):292.

36. Abagnale L, Candia C, Motta A, Galloway B, Ambrosino P, Molino A, Maniscalco M. Flow-mediated dilation as a marker of endothelial dysfunction in pulmonary diseases: A narrative review. Respiratory medicine and research. 2023;84(101049):101049.

37. Boivin DB, Boudreau P. Impacts of shift work on sleep and circadian rhythms. Pathologie-biologie. 2014;62(5):292–301.

38. Vasconcelos SP, Lemos LC, Moreno CRC. Night shift work and sleep disturbances in women: A scoping review. Sleep medicine clinics. 2023;18(4):533–543.

39. Csoma B, Bikov A. The role of the circadian rhythm in dyslipidaemia and vascular inflammation leading to atherosclerosis. International journal of molecular sciences. 2023;24(18).

40. Poza JJ, Pujol M, Ortega-Albás JJ, Romero O, Insomnia Study Group of the Spanish Sleep Society (SES). Melatonin in sleep disorders. Neurología (English Edition). 2022;37(7):575–585.

41. Zhang H-M, Zhang Y. Melatonin: a well-documented antioxidant with conditional pro-oxidant actions. Journal of pineal research. 2014;57(2):131–146.

42. Hoseini SG, Heshmat-Ghahdarijani K, Khosrawi S, Garakyaraghi M, Shafie D, Roohafza H, Mansourian M, Azizi E, Gheisari Y, Sadeghi M. Effect of melatonin supplementation on endothelial function in heart failure with reduced ejection fraction: A randomized, double-blinded clinical trial. Clinical cardiology. 2021;44(9):1263–1271.

43. Moriguchi J, Itoh H, Harada S, Takeda K, Hatta T, Nakata T, Sasaki S. Low frequency regular exercise improves flow-mediated dilatation of subjects with mild hypertension. Hypertension research: official journal of the Japanese Society of Hypertension. 2005;28(4):315–321.

44. Davoodi M, Hesamabadi BK, Ariabood E, Izadi MR, Ghardashi-Afousi A, Bigi MAB, Asvadi-Fard M, Gaeini AA. Improved blood pressure and flow-mediated dilatation via increased plasma adropin and nitrate/nitrite induced by high-intensity interval training in patients with type 2 diabetes. Experimental physiology. 2022;107(8):813–824.

45. Ades PA, Savage PD, Lischke S, Toth MJ, Harvey-Berino J, Bunn JY, Ludlow M, Schneider DJ. The effect of weight loss and exercise training on flow-mediated dilatation in coronary heart disease: a randomized trial. Chest. 2011;140(6):1420–1427.

